# Patients’ Experiences in a Dialogue-Based Outpatient Clinic Embedded Within Routine Oncology Care

**DOI:** 10.64898/2026.10.01.26364357

**Authors:** Keiko Yoshimura, Sae Fujioka, Paul Matthew Lewis, Mayuko Hirata, Misato Tani, Kohei Akazawa, Norio Kohno

## Abstract

**Objectives:** Contemporary oncology has achieved remarkable advances in biomedical treatment while increasingly emphasizing patient-centered care. Nevertheless, routine oncology consultations must address multiple competing clinical priorities, potentially limiting opportunities for patients’ experiences to become visible. To address this gap, we established a Dialogue-Based Outpatient Clinic, a complementary patient-directed consultation model embedded within routine oncology care. This study explored the dimensions of patients’ experiences that became visible through this consultation model.

**Methods:** We conducted a qualitative study of 44 consecutive patients who attended a Dialogue-Based Outpatient Clinic embedded within routine oncology care between 2021 and 2023. Patients attended the clinic either on their own initiative or following a recommendation from a healthcare professional who recognized that they were struggling. Regardless of how they came to the clinic, patients independently determined the consultation agenda.

**Results:** Of the 44 patients, 13 (29.5%) sought biomedical information regarding diagnosis, treatment, or adverse effects, whereas 31 (70.5%) discussed concerns extending beyond biomedical issues. Reflexive thematic analysis of these 31 patients identified four interconnected themes: (1) making sense of a cancer diagnosis within one’s own life; (2) negotiating treatment in relation to personal values and life priorities; (3) being treated for disease but not seen as a person; and (4) reconstructing life and identity through the cancer journey. Across all themes, patient-directed dialogue consistently revealed how patients understood cancer within the broader context of their lives rather than as an isolated biomedical condition.

**Conclusions:** Patient-directed dialogue within routine oncology care revealed dimensions of patients’ experiences that may be less readily recognized in conventional consultations. A complementary consultation model that allows patients to determine the agenda may provide opportunities for these experiences to become clinically recognizable.

Practice implications: Integrating patient-directed consultations into routine oncology care may provide clinicians with opportunities to recognize and address patients’ concerns that might otherwise remain outside the usual consultation agenda.

## Introduction

Contemporary oncology has achieved remarkable advances in improving outcomes for people with cancer. Alongside these advances, the field has increasingly embraced patient-centered care through improvements in communication, shared decision-making, advance care planning, survivorship care, and supportive care.1–4 Consequently, contemporary oncology seeks not only to optimize biomedical outcomes but also to understand and support patients’ experiences throughout the cancer journey.

Routine oncology consultations must address multiple competing priorities and often focus on immediate clinical concerns, potentially limiting opportunities for patients’ experiences to become visible within routine oncology care. Previous studies have substantially advanced understanding of patients’ experiences through questionnaires, interviews, focus groups, and observational research.2,5 Despite these important advances, it remains unclear what aspects of patients’ experiences become visible when patients are free to determine the consultation agenda within an ongoing clinical service.

To address this question, we established a physician-led Dialogue-Based Outpatient Clinic embedded within routine oncology care, in which patients freely determined the consultation agenda. Using this patient-directed consultation model, we explored which dimensions of patients’ experiences became visible within routine oncology care.

## Methods

### Study Design

This qualitative study used reflexive thematic analysis following the approach described by Braun and Clarke, with themes developed inductively through engagement with the consultation records.5,6 The study explored which dimensions of patients’ experiences became visible within the Dialogue-Based Outpatient Clinic (“Ohanashi Clinic”). The Japanese word ohanashi means “dialogue.”

This study was reported in accordance with the Standards for Reporting Qualitative Research (SRQR). 7

### Study Setting

The Dialogue-Based Outpatient Clinic was established in Japan as a physician-led complementary outpatient consultation service embedded within routine oncology care. Unlike conventional oncology consultations, the clinic was designed to allow patients to attend on their own initiative or following a healthcare professional’s recommendation and to independently determine the consultation agenda without predefined topics or consultation goals. Consultations were conducted face-to-face by a physician with clinical experience in surgery, oncology, and palliative care.

### Participants

Patients who attended the Dialogue-Based Outpatient Clinic between January 2021 and December 2023 were eligible for inclusion. All participants were adults with breast cancer or undergoing diagnostic evaluation for suspected breast cancer. They attended the clinic either on their own initiative or following a recommendation from a healthcare professional who recognized that they were struggling. Regardless of how they came to the clinic, patients independently determined the consultation agenda. All 44 consecutive eligible patients were included in the analysis.

### Data Collection

Data consisted of routine electronic clinical records and, when the content of the consultation could not be adequately captured in the electronic clinical record, supplementary records prepared by the consulting physician, both documented during or after the consultations. These records documented the concerns raised by patients, relevant clinical context, aspects of the dialogue, clinical considerations, and subsequent responses or actions. Because the consultations were conducted as part of routine clinical practice rather than as research interviews, they were not audio-recorded, and verbatim transcripts were not produced.

### Data Analysis

The consultation records were repeatedly read to develop familiarity with the data. Initial codes were generated inductively from the consultation records and iteratively refined through comparison across cases, contributing to the development of the final themes. The researchers moved iteratively between the consultation records and emerging thematic interpretations. Throughout the analytic process, emerging interpretations and themes were discussed among the authors and iteratively refined while remaining grounded in the original consultation narratives and their clinical context.

During the analytic process, patients were distinguished between those whose concerns primarily involved biomedical information and those whose concerns extended beyond biomedical issues. The thematic analysis then focused on the latter group to examine the dimensions of patients’ experiences that became visible when patients were free to determine the consultation agenda. The four themes represented interconnected patterns of meaning rather than mutually exclusive categories of patients.

### Ethics

The study was approved by the Institutional Review Board of Osaka Saiseikai Nakatsu Hospital (approval numbers 2022-65 and 2024-01). Information about the study was publicly disclosed, and participants could decline participation through an opt-out process. This study was conducted in accordance with the principles of the Declaration of Helsinki. 10

## Results

### Participant Characteristics

Between January 2021 and December 2023, 44 patients attended the Dialogue-Based Outpatient Clinic, accounting for 111 consultations. All participants were women attending breast oncology care, including one who attended before receiving a definitive diagnosis. Their mean age was 60.0 ± 13.0 years (range, 37–86 years). Participants attended a median of 1 consultation (IQR, 1–2), although some returned repeatedly throughout the cancer journey (range, 1–31 consultations). Participant characteristics are summarized in Table 1.

**Table 1.**
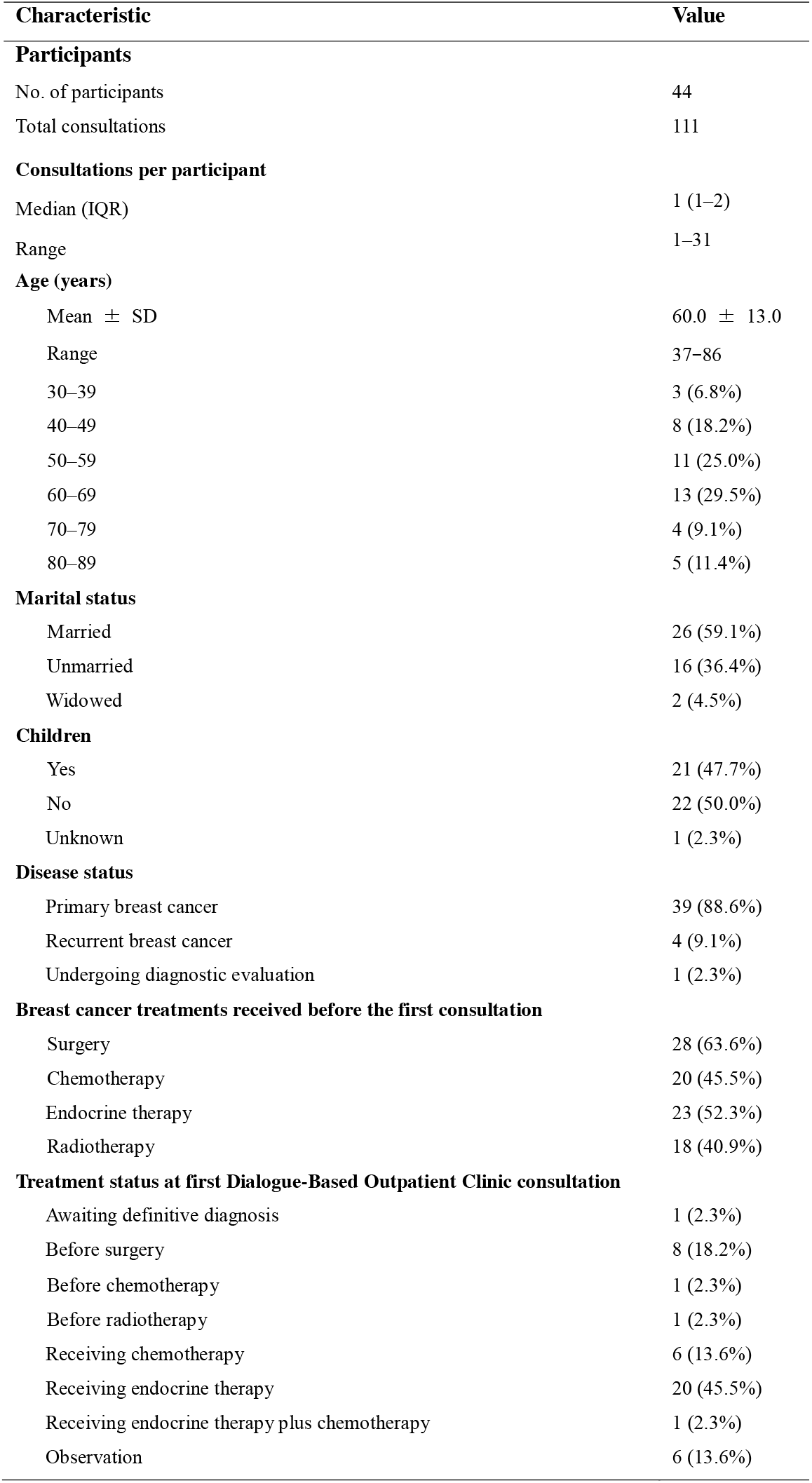
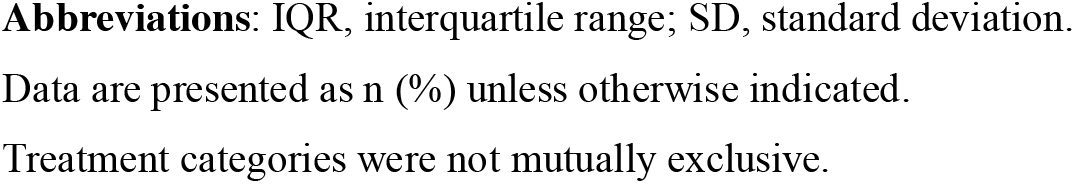
Baseline Characteristics of Participants (N = 44)

| Characteristic | Value |
| --- | --- |
| **Participants** |  |
| No. of participants | 44 |
| Total consultations | 111 |
| **Consultations per participant** |  |
| Median (IQR) | 1 (1–2) |
| Range | 1–31 |
| **Age (years)** |  |
| Mean ± SD | 60.0 ± 13.0 |
| Range | 37–86 |
| 30–39 | 3 (6.8%) |
| 40–49 | 8 (18.2%) |
| 50–59 | 11 (25.0%) |
| 60–69 | 13 (29.5%) |
| 70–79 | 4 (9.1%) |
| 80–89 | 5 (11.4%) |
| **Marital status** |  |
| Married | 26 (59.1%) |
| Unmarried | 16 (36.4%) |
| Widowed | 2 (4.5%) |
| **Children** |  |
| Yes | 21 (47.7%) |
| No | 22 (50.0%) |
| Unknown | 1 (2.3%) |
| **Disease status** |  |
| Primary breast cancer | 39 (88.6%) |
| Recurrent breast cancer | 4 (9.1%) |
| Undergoing diagnostic evaluation | 1 (2.3%) |
| **Breast cancer treatments received before the first consultation** |  |
| Surgery | 28 (63.6%) |
| Chemotherapy | 20 (45.5%) |
| Endocrine therapy | 23 (52.3%) |
| Radiotherapy | 18 (40.9%) |
| **Treatment status at first Dialogue-Based Outpatient Clinic consultation** |  |
| Awaiting definitive diagnosis | 1 (2.3%) |
| Before surgery | 8 (18.2%) |
| Before chemotherapy | 1 (2.3%) |
| Before radiotherapy | 1 (2.3%) |
| Receiving chemotherapy | 6 (13.6%) |
| Receiving endocrine therapy | 20 (45.5%) |
| Receiving endocrine therapy plus chemotherapy | 1 (2.3%) |
| Observation | 6 (13.6%) |

**Abbreviations:** IQR, interquartile range; SD, standard deviation.
Data are presented as n (%) unless otherwise indicated.
Treatment categories were not mutually exclusive.

### Consultation Topics

Patients independently determined the consultation agenda without restrictions on the topics discussed. Of the 44 patients, 13 (29.5%) primarily sought consultation regarding biomedical questions, including diagnosis, treatment options, adverse effects, and other medical concerns. In contrast, 31 patients (70.5%) discussed concerns extending beyond biomedical issues. Rather than seeking additional medical information alone, these patients used the consultations to understand diagnosis, treatment, and medical care within the broader context of their lives. The concerns extending beyond biomedical issues were organized into four themes, which are summarized in Table 2.

**Table 2.**
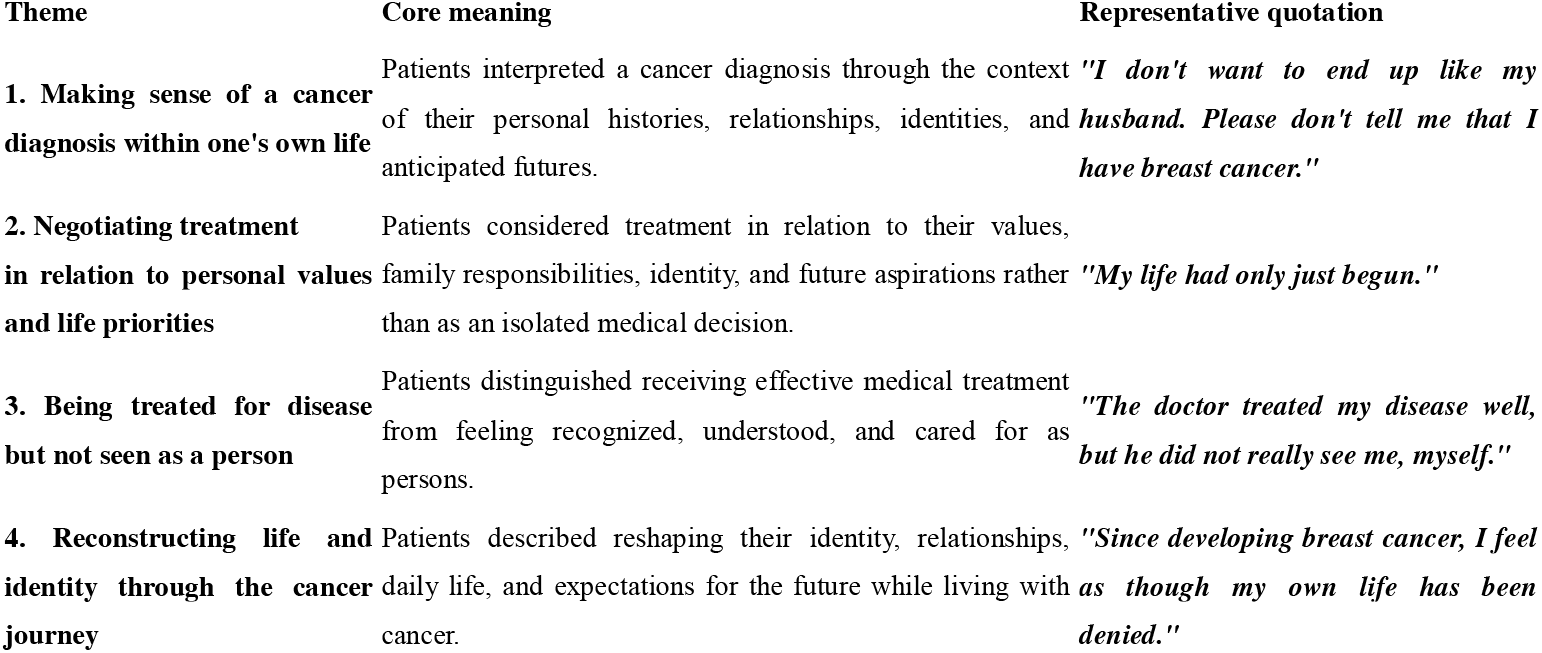
Themes and Representative Quotations from Patient-Directed Dialogue.

| Theme | Core meaning | Representative quotation |
| --- | --- | --- |
| **1. Making sense of a cancer diagnosis within one's own life** | Patients interpreted a cancer diagnosis through the context of their personal histories, relationships, identities, and anticipated futures. | *"I don't want to end up like my husband. Please don't tell me that I have breast cancer."* |
| **2. Negotiating treatment in relation to personal values and life priorities** | Patients considered treatment in relation to their values, family responsibilities, identity, and future aspirations rather than as an isolated medical decision. | *"My life had only just begun."* |
| **3. Being treated for disease but not seen as a person** | Patients distinguished receiving effective medical treatment from feeling recognized, understood, and cared for as persons. | *"The doctor treated my disease well, but he did not really see me, myself."* |
| **4. Reconstructing life and identity through the cancer journey** | Patients described reshaping their identity, relationships, daily life, and expectations for the future while living with cancer. | *"Since developing breast cancer, I feel as though my own life has been denied."* |

### Theme 1. Making Sense of a Cancer Diagnosis Within One’s Own Life

Patients discussed diagnosis not as an isolated biomedical event but in relation to their own lives. They made sense of their diagnosis in relation to previous illness experiences, family relationships, personal identity, and anticipated futures.

An older woman attended the clinic before receiving a definitive diagnosis after being informed that she had a breast tumor requiring further investigation. Several years earlier, her husband had undergone surgery for a brain tumor, never regained consciousness, and subsequently died. Before receiving a definitive diagnosis, she had already interpreted the possibility of breast cancer through the lens of her husband’s fatal illness. She understood her husband’s brain tumor and her own possible breast cancer as different forms of the same growth. Anticipating that she too might have cancer, she repeatedly said,

“I don’t want to end up like my husband. Please don’t tell me that I have breast cancer.”

Her concerns about diagnosis were inseparable from memories of her husband’s illness and death.

Overall, patients interpreted diagnosis within the context of their lives rather than as an isolated biomedical event.

### Theme 2. Negotiating Treatment in Relation to Personal Values and Life Priorities

Patients did not consider treatment in isolation from the lives they wished to live. Rather, they sought to reconcile recommended treatment with their own values, life priorities, family responsibilities, identity, and future aspirations.

A woman had devoted many years to caring for her son with a disability. After he became independent, she described entering a new stage of life in which she was free to focus on herself. She reflected,

“My life had only just begun.”

Shortly afterward, she was diagnosed with breast cancer and underwent surgery. Although she understood that guideline-recommended radiotherapy and endocrine therapy were expected to reduce the risk of recurrence, she questioned how further treatment could be reconciled with the life she had only just begun to rediscover. Through dialogue, she explored these concerns and ultimately decided to proceed with the recommended treatment.

These narratives illustrate how patients interpreted treatment decisions in relation to the lives they wished to live.

### Theme 3. Being Treated for Disease but Not Seen as a Person

Many patients acknowledged the effectiveness of contemporary oncology while simultaneously describing experiences of not feeling recognized or understood as individuals.

One woman continued to experience profound emotional distress despite completing guideline-recommended treatment and achieving an excellent biomedical outcome at another institution. Because her distress persisted, she was referred for psychiatric assessment. Reflecting on this experience during the consultation, she explained that her suffering had not first been explored in relation to what she had experienced and lost throughout cancer treatment. She said,

“The doctor treated my disease well, but he did not really see me, myself.”

Across these consultations, patients described how successful disease treatment alone did not necessarily mean being seen, understood, and cared for as persons.

### Theme 4. Reconstructing Life and Identity Through the Cancer Journey

Patients described cancer not as an isolated episode but as an experience involving the continual reconstruction of identity, relationships, everyday life, and future expectations.

One woman reflected,

“Since developing breast cancer, I feel as though my own life has been denied.”

Others described reassessing family relationships, work, personal identity, and future aspirations while learning to integrate cancer into their lives.

Patients consistently described an ongoing process of reconstructing identity, relationships, and future lives throughout the cancer journey.

### Overall Characteristics of Patient-Directed Consultations

Although some patients raised biomedical questions, most brought concerns that extended beyond biomedical issues as they sought to integrate cancer into the broader context of their lives. Across all four themes, diagnosis, treatment, medical care, and survivorship were understood within the broader context of patients’ identities, relationships, values, and future lives rather than as isolated biomedical events.

Taken together, these findings showed that when patients were free to determine the consultation agenda, dimensions of patients’ experiences that may be less readily recognized during routine oncology consultations became visible.

## Discussion

This qualitative study found that a physician-led, patient-directed Dialogue-Based Outpatient Clinic embedded within routine oncology care enabled dimensions of patients’ experiences to become clinically visible. Although some patients sought consultation regarding biomedical questions, most brought concerns that extended beyond biomedical issues, understanding diagnosis, treatment, medical care, and survivorship within the broader context of their personal histories, relationships, values, and future lives. These findings suggest that a physician-led, patient-directed consultation model can broaden what becomes clinically visible within routine oncology care.

The principal contribution of this study lies not only in describing patients’ experiences but also in illustrating how a dialogue-based consultation model embedded within routine oncology care enabled these experiences to emerge within clinical practice. Unlike conventional oncology consultations, in which consultation agendas are largely shaped by biomedical priorities, patients in this clinic independently determined what they wished to discuss, whether they attended on their own initiative or following a healthcare professional’s recommendation. Consequently, concerns extending beyond biomedical issues emerged naturally within routine clinical care. This consultation model therefore provides a practical approach to identifying dimensions of patients’ experiences that might otherwise remain unrecognized despite appropriate biomedical management.

The Dialogue-Based Outpatient Clinic was intended to complement rather than replace routine oncology care. Contemporary oncology appropriately prioritizes diagnosis, treatment, and surveillance while addressing multiple competing clinical demands. Within this context, patient-directed dialogue may extend the capacity of routine oncology care to recognize dimensions of patients’ experiences that are less readily recognized during conventional consultations, thereby supporting a more patient-centered approach to oncology care.

Our findings also resonate with Arthur Kleinman’s distinction between disease and illness.8 Although contemporary oncology has achieved remarkable success in diagnosing and treating disease, patients in this study consistently understood diagnosis, treatment, and survivorship within the broader contexts of their identities, relationships, values, and future lives. The Dialogue-Based Outpatient Clinic created a clinical setting in which these illness experiences could emerge alongside biomedical concerns. These findings suggest that patient-directed dialogue complements routine oncology care by creating opportunities for illness experiences to become clinically recognizable alongside biomedical concerns.

Taken together, these findings suggest that the issue is not simply communication but what can become recognizable within routine oncology care. In this sense, patient-directed dialogue is not simply a communication technique but a clinical approach that broadens what can be recognized within routine oncology care. This perspective is also compatible with longstanding discussions regarding reductionism in medicine.9 Biomedical reductionism has been indispensable to the extraordinary scientific progress of modern oncology by enabling increasingly precise diagnosis and treatment. At the same time, biomedical categories alone cannot fully encompass the complexity of patients’ lived experiences. Rather than opposing biomedical care, patient-directed dialogue may complement routine oncology care by broadening what can be recognized in everyday clinical practice.

This study has several limitations. It was conducted at a single institution in Japan. All participants were women attending breast oncology care, including one who attended before receiving a definitive diagnosis. Consultations were not audio-recorded, and verbatim transcripts were not produced. Analysis relied on routine electronic clinical records and supplementary records prepared when needed during or after consultations. Consequently, the analysis depended on what was documented rather than on complete records of the dialogue. The identified themes represent one possible reflexive interpretation of these consultation narratives; alternative interpretations are also possible, as expected in reflexive qualitative research. The transferability of these findings to other clinical settings and patient population requires further investigation.

Future research should examine implementation across diverse oncology settings and evaluate whether this consultation model improves recognition of patients’ experiences and influences communication, shared decision-making, survivorship care, advance care planning, patient-reported outcomes, patient satisfaction, and healthcare utilization.

## Conclusion

Patient-directed dialogue within routine oncology care revealed dimensions of patients’ experiences that may be less readily recognized in conventional consultations. Across the four themes, patients understood diagnosis, treatment, and survivorship within the broader contexts of their lives rather than as isolated biomedical events. The Dialogue-Based Outpatient Clinic provided a complementary clinical setting in which these experiences could become recognizable alongside biomedical concerns.

### Practice implications

Integrating patient-directed consultations into routine oncology care may provide clinicians with opportunities to recognize and address patients’ concerns that might otherwise remain outside the usual consultation agenda. Allowing patients to determine what to discuss, regardless of how they came to the clinic, may complement existing oncology services without replacing their biomedical functions. Further research is needed to evaluate the implementation and clinical effects of this consultation model.

## Data Availability

Data availability:
The data supporting the findings of this study are not publicly available due to the sensitive and potentially identifiable nature of the clinical narratives and the need to protect patient confidentiality.

## Declarations

## Ethics approval

The study was approved by the Institutional Review Board of Osaka Saiseikai Nakatsu Hospital (approval numbers 2026-60).

## Data availability

The data supporting the findings of this study are not publicly available due to the sensitive and potentially identifiable nature of the clinical narratives and the need to protect patient confidentiality.

## Author contributions (CRediT)

Keiko Yoshimura: Conceptualization, Methodology, Investigation, Data curation, Formal analysis, Writing – original draft, Writing – review & editing, Supervision, Project administration. Sae Fujioka, Paul Matthew Lewis, Mayuko Hirata, Misato Tani, Kohei Akazawa, Norio Kohno: Conceptualization, Formal analysis, Writing – review & editing.

## Acknowledgements

We sincerely thank all the patients who generously shared their experiences through the Dialogue-Based Outpatient Clinic. We also thank the clinicians and staff of the Department of Breast Surgery at Osaka Saiseikai Nakatsu Hospital for their support in establishing and conducting this clinic. In particular, we are grateful to Yurika Machida, Ayuka Tanaka, Yurie Suenaga, Miku Oishi, and Yumi Sawada for their help.

## Figure legend

**Figure 1.**
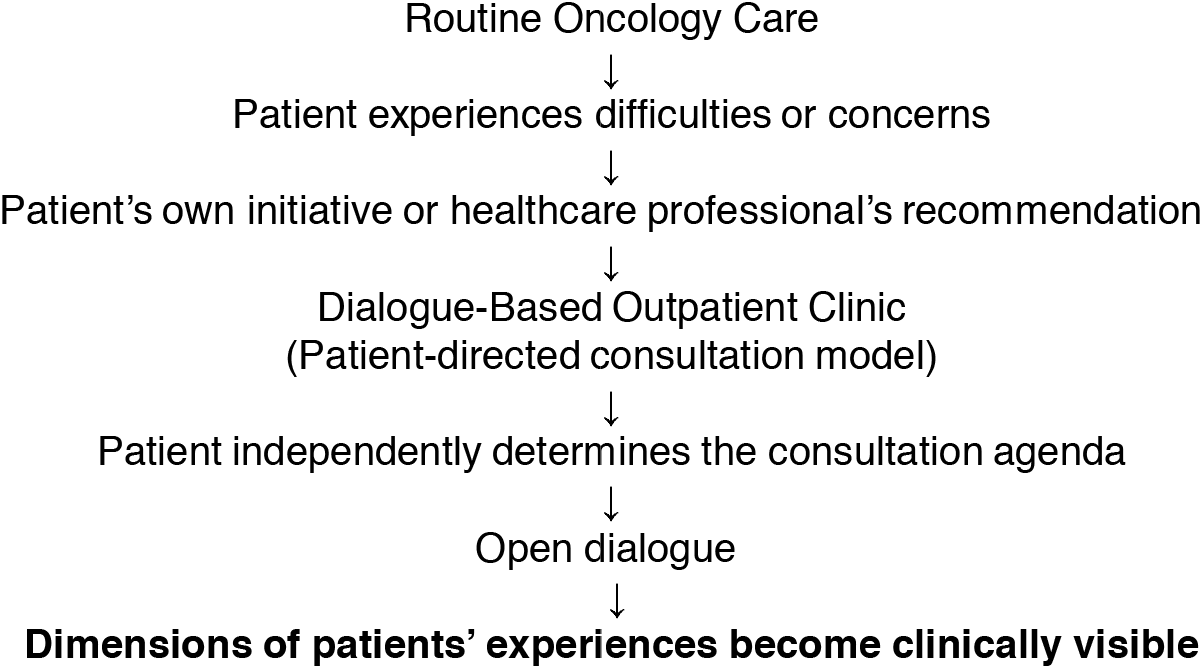
Conceptual model of the Dialogue-Based Outpatient Clinic.

The Dialogue-Based Outpatient Clinic was embedded within routine oncology care as a complementary patient-directed consultation service. Patients attended on their own initiative or following a healthcare professional’s recommendation and independently determined the consultation agenda. Through open dialogue, dimensions of patients’ experiences that might otherwise remain unrecognized entered clinical attention, broadening what became visible within routine oncology care.

